# Detection of neutralising antibodies to SARS coronavirus 2 to determine population exposure in Scottish blood donors between March and May 2020

**DOI:** 10.1101/2020.04.13.20060467

**Authors:** CP. Thompson, N. Grayson, R.S. Paton, J. Bolton, J. Lourenco, BS. Penman, L. Lee, V. Odon, J. Mongkolsapaya, S. Chinnakannan, W. Dejnirattisai, M. Edmans, A. Fyfe, C. Imlach, K. Kooblall, N. Lim, C. Liu, C. Lopez-Camacho, C. McInally, A.L. McNaughton, N. Ramamurthy, J. Ratcliff, P. Supasa, O. Sampson, B. Wang, A. Mentzer, M. Turner, M.G. Semple, J.K. Baillie, ISARIC4C Investigators, H. Harvala, G.R. Screaton, N. Temperton, P. Klenerman, L.M. Jarvis, S. Gupta, P. Simmonds

## Abstract

**Background:** The progression and geographical distribution of SARS coronavirus 2 (SARS-CoV-2) infection in the UK and elsewhere is unknown because typically only symptomatic individuals are diagnosed. We performed a serological study of blood donors in Scotland between the 17^th^ of March and the 18^th^ of May to detect neutralising antibodies to SARS-CoV-2 as a marker of past infection and epidemic progression.

**Aim:** To determine if sera from blood bank donors can be used to track the emergence and progression of the SARS-CoV-2 epidemic.

**Methods:** A pseudotyped SARS-CoV-2 virus microneutralisation assay was used to detect neutralising antibodies to SARS-CoV-2. The study group comprised samples from 3,500 blood donors collected in Scotland between the 17^th^ of March and 19^th^ of May, 2020. Controls were collected from 100 donors in Scotland during 2019.

**Results:** All samples collected on the 17^th^ March, 2020 (n=500) were negative in the pseudotyped SARS-CoV-2 virus microneutralisation assay. Neutralising antibodies were detected in 6/500 donors from the 23^th^-26^th^ of March. The number of samples containing neutralising antibodies did not significantly rise after the 5^th^-6^th^ April until the end of the study on the 18^th^ of May. We find that infections are concentrated in certain postcodes indicating that outbreaks of infection are extremely localised. In contrast, other areas remain comparatively untouched by the epidemic.

**Conclusion:** These data indicate that sero-surveys of blood banks can serve as a useful tool for tracking the emergence and progression of an epidemic like the current SARS-CoV-2 outbreak.

## Introduction

SARS coronavirus 2 (SARS-CoV-2) emerged in late 2019 in Hubei province China as a cause of respiratory disease occasionally leading to acute respiratory distress syndrome and death (COVID-19) (1,2). On the 11^th^ of March, the WHO declared the SARS-CoV-2 outbreak a pandemic. As of July 2020, roughly 10 million confirmed cases of COVID-19 have occurred resulting in 500,000 deaths (3). Increasing age, male gender, smoking and comorbidities such as cardiac disease, hypertension and diabetes have been identified as risk factors for severe infections (4,5).

Symptomatic individuals typically exhibit fever, cough and shortness of breath 2-14 days after infection (6). However, an unknown proportion of individuals experience no symptoms (7–9). Antibody responses in both symptomatic and asymptomatic individuals are detectable in the blood 14 to 28 days after infection (10,11). Subsequently antibody levels drop and can become undetectable by some antibody assays in the early convalescent phase (10,12,13)

In this study, we follow blood donors as a means of estimating population exposure from the start of the pandemic in March through to mid-May when PCR-detected cases in the UK had plateaued (14,15). Samples from donors in an age range of 18-75 years collected across Scotland were assayed for neutralising antibody to SARS-CoV-2 using a pseudotyped SARS-CoV-2 virus microneutralisation (pMN) assay used previously for SARS-CoV and Ebola virus seroepidemiology purposes (16–18). The sensitivity of the neutralisation assay was confirmed using an enzyme-immunosorbant (ELISA) assay detecting antibodies to spike protein. The detection frequency of neutralising antibodies in blood donors and a discussion of its applicability for estimating population level exposure are presented.

## Methods

### Samples

500 plasma samples collected on the 17^th^ March, 21^st^-23^rd^ March, 5^th^-6^th^ April, 18^th^-20^th^ April, 2^nd^-4^th^ May and 16^th^-18^th^ May were analysed in the study. An additional 500 samples from the Greater Glasgow region, collected between the 2^nd^-4^th^ of May were also analysed. To serve as negative controls, 100 blood donor samples were tested in parallel from the Scottish National Blood Transfusion Service (SNBTS) anonymous archive collected between September 2018 and December 2019 (IRAS Project No. 18005), before the first reports of the spread of SARS-CoV-2 in China (1,2). Seventeen control samples from contract-traced individuals who were PCR-confirmed as SARS-CoV-2 infected were used as positive controls in the study. All the individuals from whom the positive control sera samples were taken had asymptomatic SARS-CoV-2 infections and were recruited through the ISARIC WHO Clinical Characterisation Protocol UK (CCP-UK) at the discharge plus 28 day time-point. Samples were heat inactivated prior to serological testing by incubation at 56°C for 30 minutes.

### SARS-CoV-2 pseudotype microneutralisation assay

A lentivirus-based SARS-CoV-2 pseudovirus particle was constructed displaying the full spike protein on the surface of the pseudotyped virus using a synthetic codon optimised SARS-CoV-2 expression construct (Accession number: YP_009724390.1). Virus infectivity was determined by titration on HEK 293T ACE2-plasmid transfected cells as previously described (19). Neutralising antibody titres were determined by endpoint two-fold serial dilutions of test samples mixed with 10^5^ relative light units (RLU) of pseudotyped virus, incubated at 37°C for two hours and then mixed with 10^4^ HEK 293T ACE2-transfected cells per well. Plates were incubated for 72 hours at 37°C and then cells were lysed and assayed for luciferase expression. Neutralisation titres are expressed as *IC*_50_ values. During the assay, plates were barcoded and controls were spaced throughout the runs. Individuals were blinded regarding the arrangement of spaced positive controls on the plates.

### Titration

Pre-pandemic samples and samples collected on the 17^th^ March and the 21^st^-23^rd^ March were all titrated to optimise the neutralisation assay. After this point, samples were initially screened for neutralisation using the highest 1:20 dilution. Dilutions of 1:20 were performed in triplicate along with virus-only, no virus and positive control wells. Samples that produced a mean RLU two standard deviations below the mean were then titrated out to obtain *IC*_50_values.

### Enzyme-linked immunosorbent assay (ELISA)

Antibodies to the trimeric spike protein were detected by ELISA. MAXISORP immunoplates (442404; NUNC) were coated with StrepMAB-Classic (2-1507-001;iba). Plates were blocked with 2% skimmed milk in phosphate buffered saline (PBS) for one hour and then incubated with 0.125ug of soluble SARS-CoV2 trimeric spike protein or 2% skimmed milk in PBS. After one hour, plasma was added at 1:50 dilution, followed by alkaline phosphatase (AP)-conjugated anti-human IgG (A9544; Sigma) at 1:10,000 dilution or AP-conjugated anti-human IgM (A9794; Sigma) at 1:5000 dilution. The reaction was developed by the addition of p-Nitrophenyl Phosphate (PNPP) substrate and stopped with NaOH. The absorbance was measured at 405nm after 1 hour. Further information is provided in Adams et al 2020 (20).

### Estimating the 50% inhibitory concentration

RLUs for each well were standardised against technical positive (cells and virus with no sera) and negative (cells with no sera) controls on each plate to determine a percentage neutralisation value. An average neutralisation was calculated across the two sample replicates on each plate (for each dilution). Dilution curves were fit to each sample, with the percentage neutralisation modelled as a logistic function of the dilution factor. This yielded an *IC*_50_ value for each sample where a curve could be fit; samples that showed no dilution response because of complete or no neutralisation were not given an *IC*_50_ value. Dilution curves were estimated using nonlinear least squares in R version 3.6.3 (21). An error weighted mean of the *IC*_50_ value was calculated for samples that were repeated on more than one plate. We classified positive samples as having an *IC*_50_ value greater than the largest negative control (1:69) with a standard error less than or equal to the least neutralising positive control.

### Determining test sensitivity and specificity

Test sensitivity (probability of neutralisation given positive sera) and specificity (probability of a negative result given no exposure) was estimated using 17 (RT-PCR confirmed) positive controls and 100 pre-pandemic blood donor samples as negative controls. The highest inhibitory concentration observed for a negative control was used as a threshold to determine positive samples (giving 100% [98.10-100] specificity). Of the 17 positive controls, 16 samples neutralised with high confidence, giving an estimated sensitivity of 94.11% [79.17-99.98] (Figure 1).

**Figure 1.**
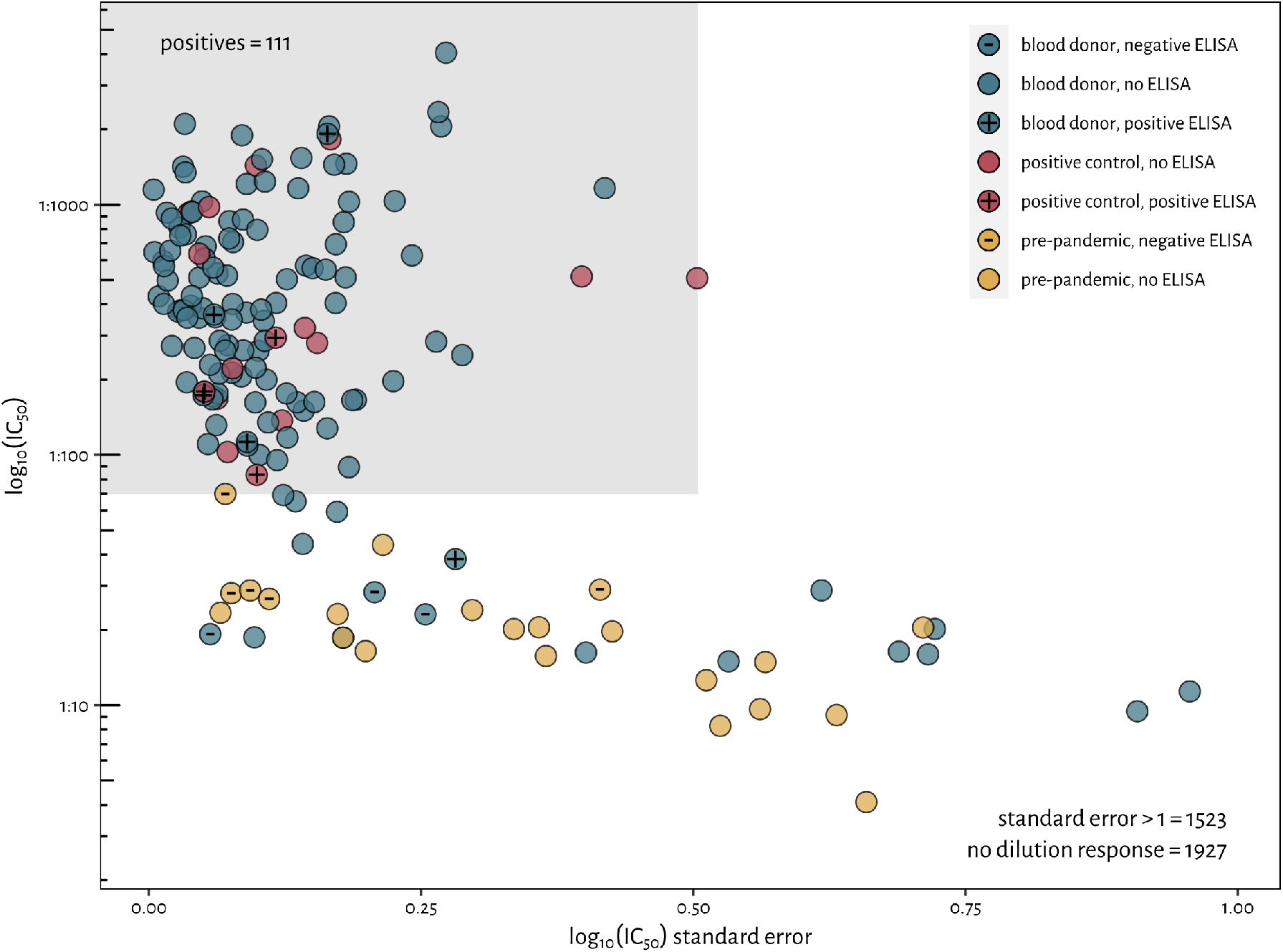
Selection criteria for classifying a sample as neutralising. Samples are required to have an estimated *IC*_50_ and a standard error at least as small as the worst neutralising positive control. This penalises samples with poorly defined inhibitory concentrations. Pre--pandemic samples are shown in yellow, positive controls in red and blood donor samples as blue. ELISA results are annotated on top of the points.

### Accounting for sensitivity and specificity in sample prevalence estimates

Uncertainty in test sensitivity and specificity can be propagated to sample prevalence estimates using a simple hierarchical Bayesian model (22). The number of positive tests in the positive (*n*^+^ = 16) and negative (*n*^−^ = 0) control groups was modelled as a binomial distribution:

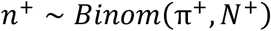

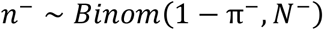

where the sensitivity is given by π^+^ and the specificity by π^−^ (*N*^+^ = 17 and *N*^−^ = 100 are the number of positive and negative controls respectively). An estimate of the true proportion of positive sera for samples from a given week and health board (*p*_*w,h*_) is comprised of neutralising sera that were missed ([1 − π^+^]) and those incorrectly identified as neutralising samples (from 1 − π^−^). The observed number of positive samples for the week, w, and health board, h, (n_*w*,h_) was modelled as a binomial distribution accounting for test performance:

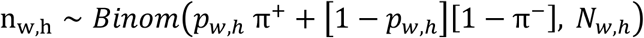

with *N*_*w,h*_ the number of samples from each health board in each week. Using this method, the uncertainty in test specificity and sensitivity is propagated to the estimate of the seroprevalence; this results in broader credible intervals that better reflect the inherent uncertainty in test parameters.

### Modelling sample prevalence

In estimating seroprevalence, we assume that neutralising antibodies do not wane in the blood donor population during the survey period and accrue to an equilibrium (13). Making this assumption, we can fit the logistic function to the time series of sample seroprevalence:

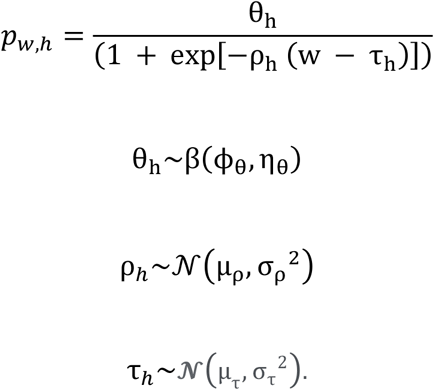

Here, *θ*_*h*_ is the equilibrium seroprevalence, ρ_h_, is the rate with which the seroprevalence approaches this maximum and τ_h_ is the midpoint of the logistic curve for each health board. Parameters were modelled using hierarchical distributions across health boards (the maximum as a beta to bound it between 0 and 1, the rate and the midpoint as a normal distribution). Priors are given in the supplementary material. The model was fit in R version 3.6.3 using the Bayesian inference package JAGS version 4.3.0 (23). Models were run across 6 chains until convergence (potential scale reduction factor less than 1.02 and effective sample size > 10,000).

### Ethics statement

Ethical approval was obtained for the SNBTS anonymous archive - IRAS Project No. 18005. SNBTS blood donors gave fully informed consent to virological testing, donation was made under the SNBTS Blood Establishment Authorisation and the study was approved by the SNBTS Research and Sample Governance Committee.

## Results

The estimated *IC*_50_ values and standard errors for the control and blood donor samples are shown in Figure 1. Of the 3500 post-pandemic blood donor samples, a total of 111 contained anti-SARS2 neutralising antibodies using the *IC*_50_ and standard error based thresholds described in the methods. The results of the neutralisation assay were positively correlated with ELISA optical density (Figure S2, Pearson’s correlation coefficient = 0.86, p < 0.001).

No samples from the 17^th^ March showed neutralising activity. Blood donor samples obtained from donations during the 21^st^-23^rd^ March, 5^th^-6^th^ April, 18^th^-20^th^ April, 2^nd^-4^th^ May and 16^th^-18^th^ May contained neutralising anti-SARS2 antibodies (Figure 2). The number of samples containing neutralising antibodies did not significantly rise after the 5^th^-6^th^ April.

**Figure 2.**
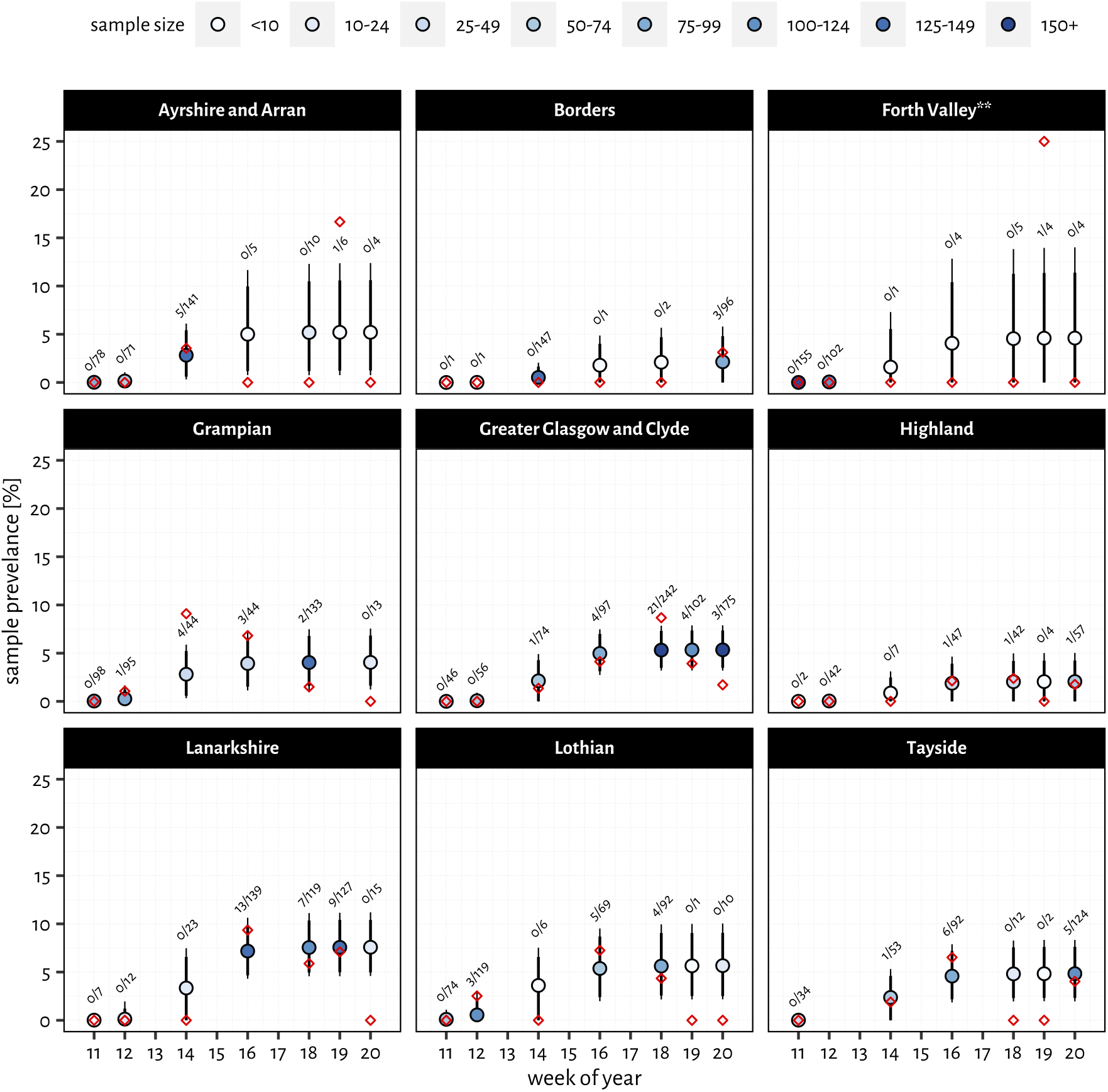
Sample prevalence estimates for each health board through time using the model outlined in the methods. Median prevalence estimates are marked with filled circles (colour denoting sample size) while thick and thin error bars give the 90 --95% highest density intervals respectively. Red diamonds represent the raw percentage estimates from the data. Estimates for Forth Valley are highlighted (**) as the poor sample coverage and single positive result could be fit by a range of values.

Estimates of seroprevalence in the healthcare boards based on the final sampling point between the 16^th^-18^th^ May are illustrated in Figures 3 and 4. The lowest uncertainty is associated with estimates from the Greater Glasgow and Clyde health board (5.35% [3.19-7.89%]); Tayside, Lothian and Grampian have similar median estimates with higher uncertainty. Lanarkshire is predicted to have the highest seroprevalence of all health boards (7.59% [4.60, 11.20%]) whilst the Highlands and Borders have the lowest seroprevalence of around 2.08 [0, 5.08%] and 2.16 [0, 5.85%], respectively. Throughout this period, *IC*_*50*_ values between weeks did not show a statistically significant difference (Figure S3). No statistically significant variation in *IC*_*50*_ value was seen based on age or sex (Figure S4).

**Figure 3.**
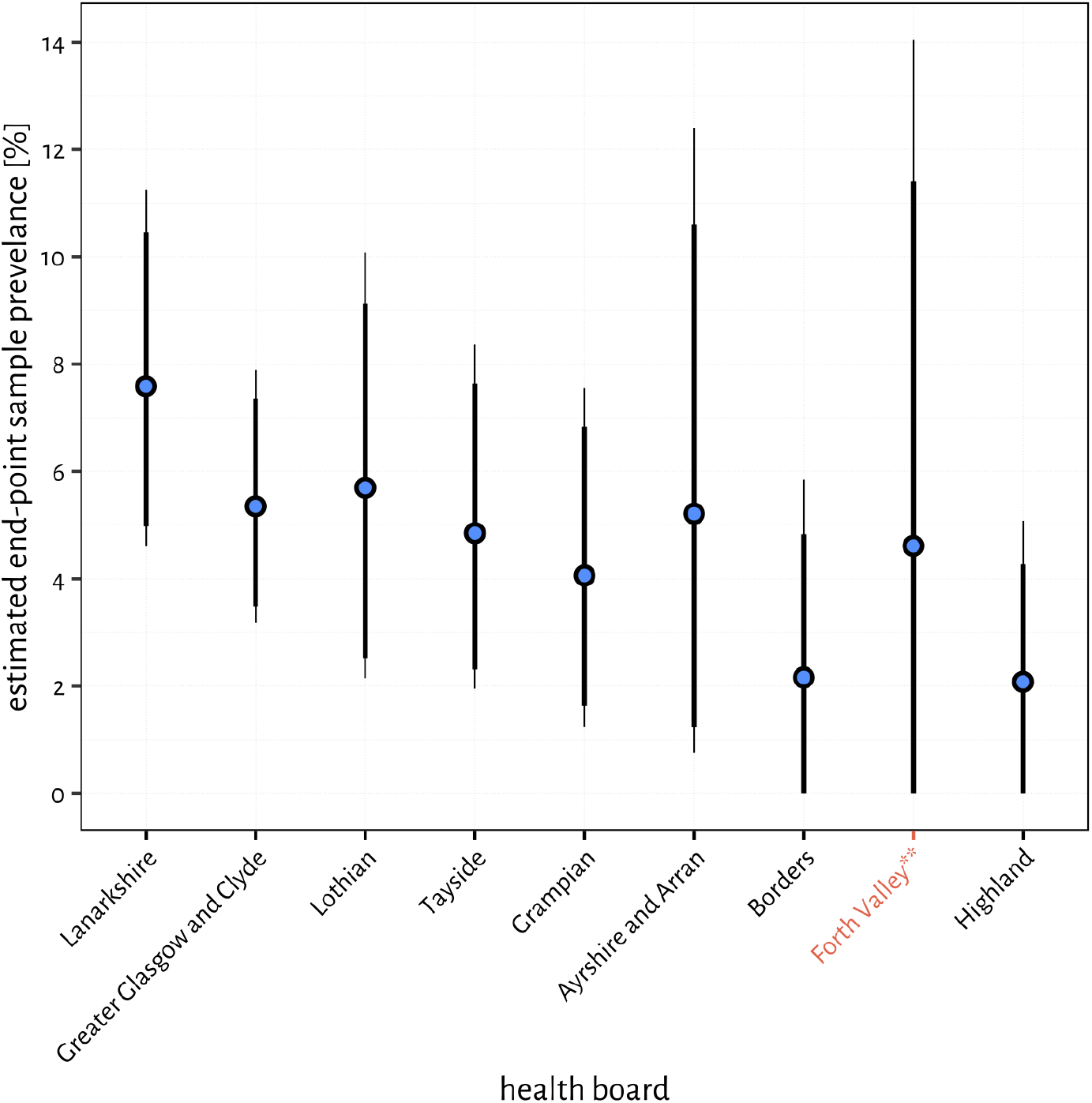
Estimates of sample prevalence at the end of our study period (the parameter *θ*_*h*_ from the logistic equation) ordered by the lower 95% confidence interval. The median parameter estimate is represented by the blue point, with thick and thin error bars denoting the 90 and 95% highest density intervals, respectively. All sample estimates are characterised by high uncertainty: the Lanarkshire health board is predicted to have the highest prevalence; Greater Glasgow and Clyde is estimated with the most confidence; while Forth Valley, Borders and Highland could not be interpreted: and estimates for the Forth Valley should be treated with scepticism due to poor sampling (see Figure 2).

**Figure 4.**
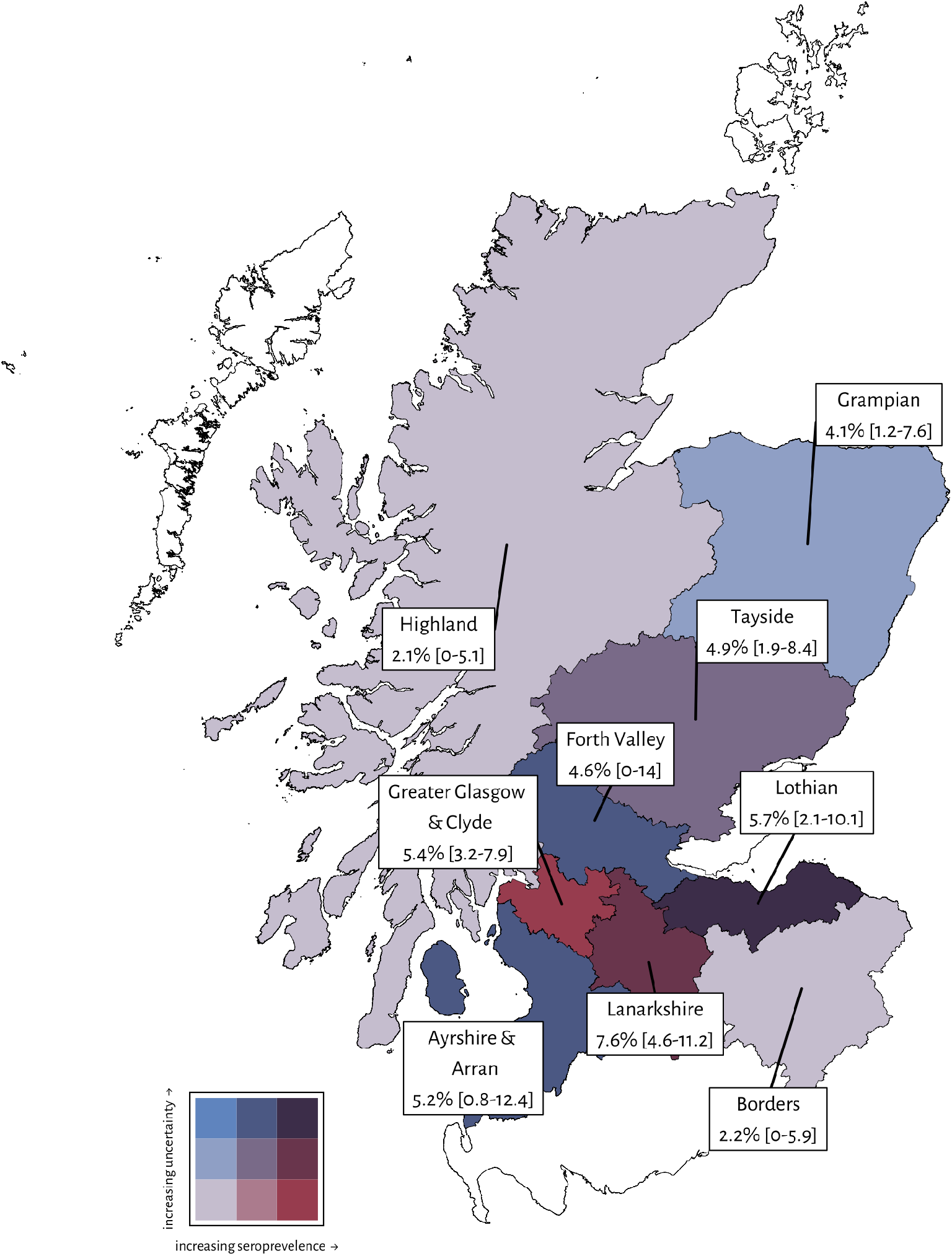
Map of Scottish health boards showing estimated endpoint seroprevalences. White health boards were not sufficiently sampled to generate estimates. The colour scale denotes the confidence and seroprevalence in each health board.

The outbreaks as a whole in Scotland were centred in the major urban centres – Glasgow and Edinburgh, in addition to the Lanarkshire health authority region (Figures 3&4). To explore this phenomenon in further detail, a separate analysis of 491 samples from the Great Glasgow region collected between the 18^th^-20^th^ April was performed. 42/490 of these samples have neutralising antibodies. Analysis of the distribution of samples containing neutralising antibodies by postcodes showed that most of these samples located in the Paisley (PA) (14/85) and Motherwell (ML) (15/197) postcodes of Greater Glasgow, indicating that outbreaks in these regions are localised. By comparison, Central Glasgow had comparatively few samples containing neutralising antibodies (7/195; Figure 5).

**Figure 5.**
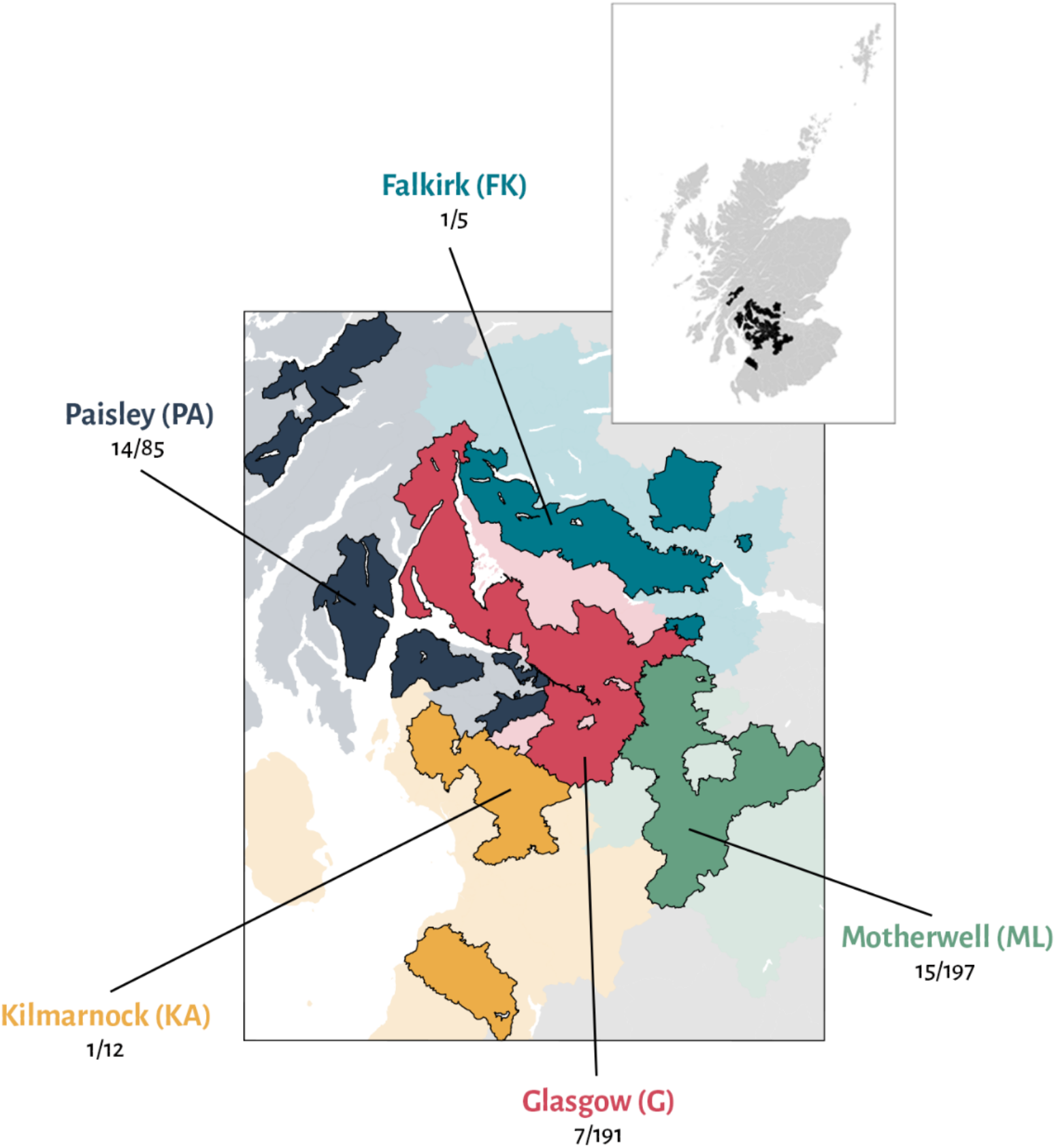
Raw counts of positive samples in the additional survey of postcodes close to Glasgow. Of the 500 samples collected, 490 fell within a reasonable radius of the city.

## Discussion

The findings from this study suggest that blood donors can be used as a sentinel population to track the emergence and progression of an epidemic.

Whilst the demographics of blood donors differs in several aspects from the general population, most notably because of the exclusion of those at risk for blood-borne viruses (HIV-1, HCV, HBV) and syphilis, they might be considered a reasonable representation of the adult population in the absence of any obvious predisposing factors for infection. The only other general exclusions were a four-week and a four month donation deferral period in those people who travelled to specified countries at risk for arbovirus and malaria infections, respectively.

However, estimates of seroprevalence are complicated by non-uniform sampling (Figures 2&3). The blood donations collected and tested in this study tend to be focused in specific postcodes, based on the locations where weekly donations took place. This produces an added level of complexity as our data shows that outbreaks are focused in specific communities even on the scale of a medium sized city such as Glasgow (Figure 5). This is further confounded by the absence of samples from individuals below the age of 18 and individuals over the age of 75.

The results presented in this study are based on a formally non-validated assay. However, by using contact traced asymptomatic individuals who had been PCR-confirmed as infected by SARS-CoV-2 and 100 donors obtained prior to the epidemic we were able to ascertain the sensitivity of the assay. Furthermore, a second ELISA based assay was used to confirm the analysis. As 16/17 PCR confirmed asymptomatic cases were detected, the assay is estimated to have a sensitivity of 94.11% [79.17-99.98] (Figure 1). Other studies have previously shown that the pMN assay correlates well with other lab-based and commercial serological assays (24).

Based on our data, the assay appears to be specific to SARS-CoV-2. There are four circulating seasonal coronaviruses: HKU1, OC43, NL63 and 229E which circulate during the winter months (25). The 100 pre-pandemic samples collected in the winter months of 2019, prior to the start of the epidemic in Scotland, did not cross-react with the SARS-CoV-2 pseudotyped virus (Figure 1). Individuals in this cohort will have been infected with the seasonal coronaviruses but not SARS-CoV-2 and contain neutralising antibodies to them.

Therefore, as there were no false positive results in the pre-pandemic samples, which were not due to noise within the neutralisation assay, we can have a high degree of confidence that the antibodies detected in the samples from March to May 2020 were generated by SARS-CoV-2 infection.

The utility of using pMN assays and ELISAs to track population exposure is dependent on the assumptions that (i) every infected individuals seroconverts, and (ii) that once seroconverted, the antibodies remain circulating in the blood at detectable levels. A decrease in total antibody and neutralsing antibody titres have been noted in samples drawn up to two months post peak neutralising antibody response (approximately 3-4 weeks post infection). In some instances antibody levels occasionally becoming undetectable when tested with a specific assay and analysis methodology (10,13). This drop in titres may lead to false negatives in the later timepoints. However, the dates of collection used in this study all fall within three months of the diagnosis of the first confirmed case in Scotland on 1^st^ March (26). For this reason, it is unlikely that this study is hampered by the drop in neutralising antibody levels described elsewhere, although future seroprevalence studies may potentially underestimate the true level of population exposure. It is possible that some individuals also may not seroconvert, representing a small pool of patients that will be false negative (12).

Samples containing anti-SARS2 neutralising antibodies were detected in blood donors who gave blood between the 16^th^-17^th^ March in all Health Boards (Figures 2&3). Subsequently, samples containing anti-SARS2 neutralising antibodies were detected in every further time point assayed until the end of the study. Consequently, due to the 14-28 day incubation period prior to seroconversion, it is likely that SARS-CoV-2 began circulating in Scotland in late February 2020 and potentially earlier (10,11).

## Data Availability

All data obtained in the study is available from the authors upon request

## Conflict of interest statement

The authors declare no conflict of interest

## Funding statement

This work was supported by the Georg & Emily Opel Foundation. This work was supported by the Medical Research Council [grant number MC_PC_19059]. National Institute for Health Research Biomedical Research Centre Funding Scheme (to G.R.S.), the Chinese Academy of Medical Sciences (CAMS) Innovation Fund for Medical Science (CIFMS), China (grant number: 2018-I2M-2-002). PK, PS and G.R.S. is supported as a Wellcome Trust Senior Investigator (grant 095541/A/11/Z; WT109965/MA). PK is an NIHR Senior Investigator. NG was supported via grant to Philip Goulder (WTIA Grant WT104748MA) and a grant to John Frater (Medical Research Council MR/L006588/1). CPT was funded by an ERC research grant ‘UNIFLUVAC’ and two MRC CiC grants (Ref: BR00140). LNL is supported by a CRUK Cancer Immunology Award (C30332/A23521) to PK.The funders played no role in the design, execution or reporting of the study. RP also supported by funds provided under Professor RW Snow’s Wellcome Trust Principal Fellowship (# 212176). ALM is funded by a NIHR Research Capability Funding grant.

MG Semple, P Klenerman, and P Simmonds are affiliated to the National Institute for Health Research Health Protection Research Unit (NIHR HPRU) in Emerging and Zoonotic Infections at University of Liverpool in partnership with Public Health England (PHE), in collaboration with Liverpool School of Tropical Medicine and the University of Oxford [award number NIHR200907]. The work was also supported by the NIHR Biomedical Research Centre, Oxford. The views expressed are those of the author(s) and not necessarily those of the MRC, NHS, the NIHR, the Department of Health or Public Health England.

## Acknowledgements

We would like to acknowledge the help and collaboration of many SNBTS staff for provision and preparation of samples (anonymous archive and recent donation samples). We acknowledge the wider support of ISARIC4C.

## Author’s Contribution

Thompson, C., Chinnakannan, S., Dejnirattisai, W., Edmans, M., Fyfe, A., Kooblall, K., Lee, L., Lim, N., Liu, C., López-Camacho, C., Mongkolsapaya, J., Odon, V., Sampson, O., Ramamurthy, N., Ratcliff, J., Supasa, P., Wang, B. and Mentzer, A., performed the sample acquisition, laboratory testing and reporting of the pseudotype and ELISA testing. Imlach C., McInally C., Harvala, H. and Jarvis, L.M. established the sample sets, archiving and data provision of the samples used in the study. Grayson, N., Lourenco, J., Penman, B.S., Semple, M.G., Baillie JK, Bolton, J. and Paton, R.S. performed the data analysis and results interpretation. Paton, R.S, Gupta., Thompson, C., Lourenco, J., wrote and interpreted the seroprevalence model. Turner, M., Thompson, C., Temperton, N., Gupta, S., Klenerman, P., Screaton, G.R. and Simmonds, P conceived and designed the specifics of the study, the data interpretation and drafting of the manuscript. All co-authors contributed to the editing and final drafting of the manuscript and figures.

## Notes

### Competing Interest Statement

The authors have declared no competing interest.

